# Causal Analysis for Multivariate Integrated Clinical and Environmental Exposures Data

**DOI:** 10.1101/2022.12.20.22283734

**Authors:** Meghamala Sinha, Perry Haaland, Ashok Krishnamurthy, Bo Lan, Stephen A. Ramsey, Patrick L. Schmitt, Priya Sharma, Hao Xu, Karamarie Fecho

## Abstract

Electronic health records (EHRs) provide a rich source of observational patient data that can be explored to infer underlying causal relationships. These causal relationships can be applied to augment medical decision-making or suggest hypotheses for healthcare research. In this study, we explored a large-scale EHR dataset on patients with asthma or related conditions (N = 14,937). The dataset included integrated data on features representing demographic factors, clinical measures, and environmental exposures. The data were accessed via a service named the Integrated Clinical and Environmental Service (ICEES). We estimated underlying causal relationships from the data to identify significant predictors of asthma attacks. We also performed simulated interventions on the inferred causal network to detect the causal effects, in terms of shifts in probability distribution for asthma attacks.

## 1 Introduction

Causal inference [1] is re-emerging as an important tool in the domain of health sciences for informatics work such as finding effects of a drug or risk factors for a disease. Causality has traditionally been a core concept across all branches of medical science and considered when diagnosing patients based on their symptoms, effects of treatment, and years of historical evidence [2]. Electronic health records (EHRs) present a potential data source to analyze digital patient information like medical history, diagnoses, medications, and laboratory results. Inferring causal relationships from these data is useful for important tasks like prediction and explanation. For prediction, we want to measure the likelihood of occurrence of an event as a result of another event, for example, the occurrence of lung cancer based on exposure to smoke in the environment. However, such predictions are susceptible to fallacies if made only based on associations for instance, an increase in the sales of matches (e.g., in a blackout-prone area) can also be associated with lung cancer. Most black-box prediction models, unlike causal inference, are not able to identify confounding variables and hence cannot differentiate causal versus spurious associations.

Another aspect of causal inference is the ability to provide an explanation for the relationship between two events. For instance, causal inference helps us to unearth why a patient is sick and diagnose them based on the underlying cause of their symptoms and other aspects of their disease. Access to EHR data is thus critical for the advancement of clinical research and practice. However, due to the many regulations that surround clinical data, while necessary to ensure patient privacy and protection of sensitive data, access to the data for research is often challenging.

In this research, we analyzed a patient-level dataset extracted from a regulatory-compliant open service called the Integrated Clinical and Environmental Exposures Service (ICEES). ICEES supports several use cases including asthma. The ICEES data are constructed by integrating clinical data elements derived from patient EHRs and environmental exposures data derived from a variety of public sources of environmental exposures data before binning or recoding the data and stripping all protected health information.

The ICEES data are then exposed via an open application programming interface (OpenAPI). For our principal application use case, we asked if there is a causal relationship between asthma attacks and the following features: sex, race, prescriptions for prednisone, diagnoses of obesity, residential proximity to a major roadway or highway, residential density, and exposure to high levels of airborne pollutants. These features were selected because published studies, including our prior work [3, 4], have recognized them to be associated to asthma attacks. We focused on an existing ICEES cohort of patients with asthma or related conditions (see [3] for details), and we considered the number of annual emergency department (ED) or inpatient visits for respiratory issues as the primary outcome measure and indicator of asthma attacks. We used the ICEES OpenAPI to extract features that might be causally related to each other and used the resultant multivariate table for causal inference modeling. Because EHR data are purely observational, we also demonstrate a way to perform simulated external intervention, given a known causal network, to help answer important questions about the effects of clinical interventions. We use subject matter expert knowledge and publication support as our ground truth to measure the correctness of our causal inference modeling. Finally, we discuss our findings, including the benefits and limitations of our causal inference model and approach.

## 2 Analysis of the Multivariate ICEES Table

We queried the ICEES OpenAPI to generate an eight-feature multivariate table. The multivariate table analysed in this work comprised data on 14,937 patients (rows represent individual patients in the asthma cohort) and eight ICEES feature variables, per patient, namely, TotalEDInpatientVisits, Sex, Race, Prednisone, Obesity, PM2.5Exposure, RoadwayExposure, and EstResidentialDensity, where TotalEDInpatientVisits is our primary outcome variable (Table 1). In Fig. 1, we plot bar charts to show comparisons of the number of TotalEDInpatientVisits among the discrete categories of each feature. We can see that the count for zero TotalEDInpatientVisits is the largest among all categories. Upon further analysis, we found that the multivariate data table extracted from the openAPI largely consisted of patients who were inactive in the year 2010. Hence, to avoid bias and reduce noise in our analysis, we removed patients who were not active in the year of interest, meaning their EHR did not indicate any healthcare usage, by applying the “Active In Year” feature as a filter to extract a multivariate table, with Active In Year = 1 to select only patients who were active in 2010. We show the bar charts for the number of ED/inpatient visits for each feature in Fig. 1. We can observe that most of patients who were active in year 2010 only visited the ED or an inpatient clinic once. We also can see there is an imbalance among the levels in some features like Prednisone, Obesity, Race, RoadwayExposure, and Pm2.5exposure.

**Table 1:** Feature variables used to generate multivariate table.

| Feature Variable | Variable Definition and Enumeration |
| --- | --- |
| Sex | Male (0), Female (1) |
| Race | Caucasian, African American, Asian, Native Hawaiian/Pacific Islander, American/Alaskan Native, Other |
| Prednisone | Common medication for asthma-like conditions (1=Yes, 0=No) |
| Obesity | Diagnostic code for obesity anytime over ‘study’ period (1=Yes, 0=No) |
| Airborne Particulate Exposure | Abbreviated herein as "PM2.5Exposure". US Environmental Protection Agency estimated maximum daily exposure to particulate matter $\leq 2.5$-microns in diameter over ‘study’ period, binned using pandas.cut |
| Roadway Exposure | Abbreviated herein as "RoadwayExposure". US Department of Transportation distance in meters from household to nearest roadway (1 = 0-49, 2 = 50-99, 3 = 100-149, 4 = 150-199, 5 = 200-249, 6 = $\geq 250$ meters) |
| Residential Density | Abbreviated herein as "EstResidentialDensity". US Census Bureau American Community Survey 2007–2011 estimated total population [block group], binned according to US Census Bureau definitions |
| Emergency Department Visits | Abbreviated herein as "TotalEDInpatientVisits". Total number ED or inpatient visits for respiratory issue(s) over the ‘study’ period (0, 1, 2, 3, ...) |

**Fig 1:**
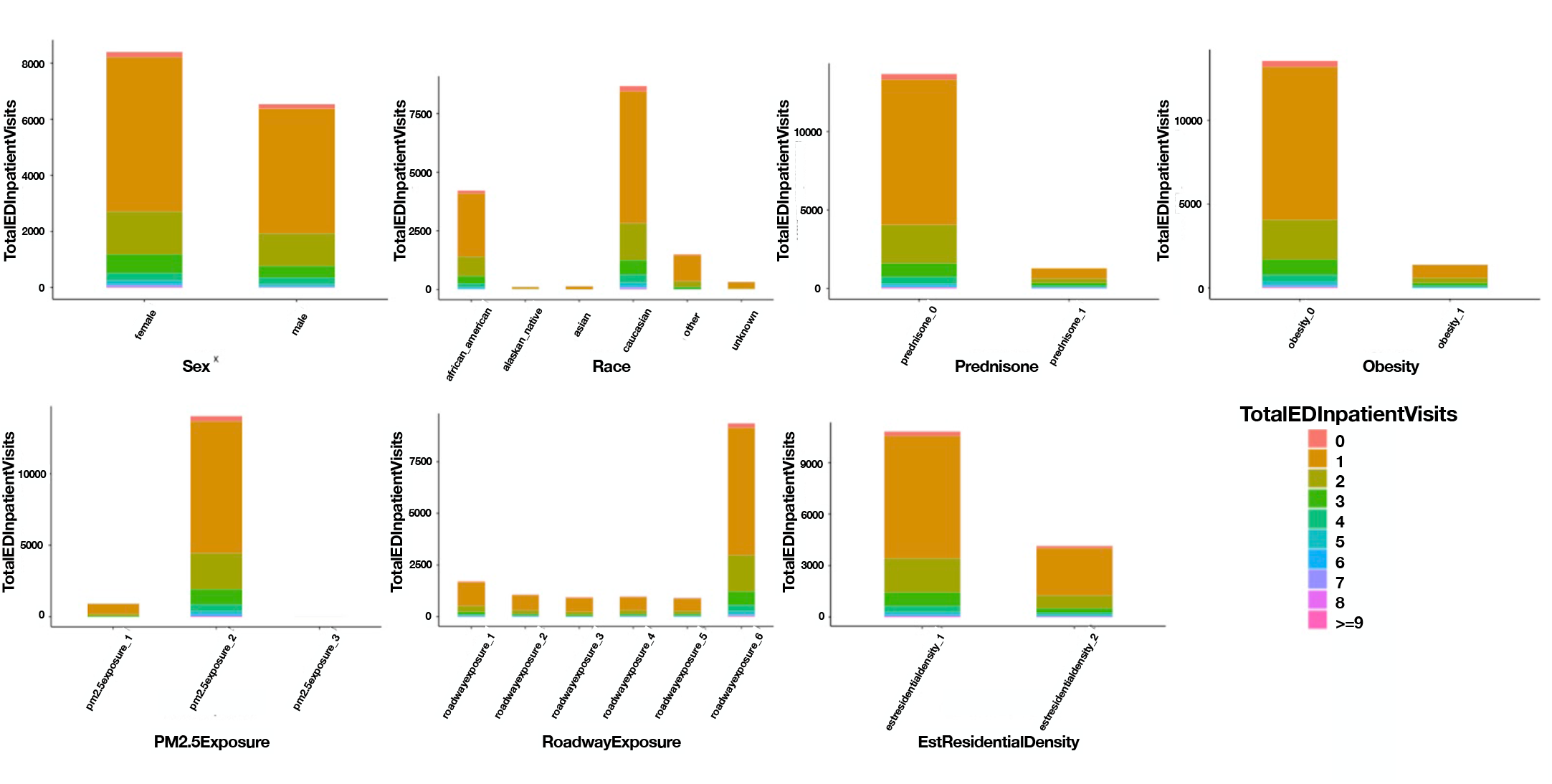
Stacked bar chart representing the number of TotalEDInpatientVisits across each level of the feature variables. See Table 1 for feature variable definitions.

### 2.1 Feature importance

We evaluated the importance of each feature and its contribution towards the model performance using a tree-based machine learning model: random forest. We leveraged the caret R package [5] to evaluate the feature importance. We controlled the parameters for training by using the *repeatedcv* method to divide our dataset into ten-folds cross-validation and repeated three times. We found Prednisone, Race, ObesityDx as the highest contributing factors, as shown in Fig 2.

**Fig 2:**
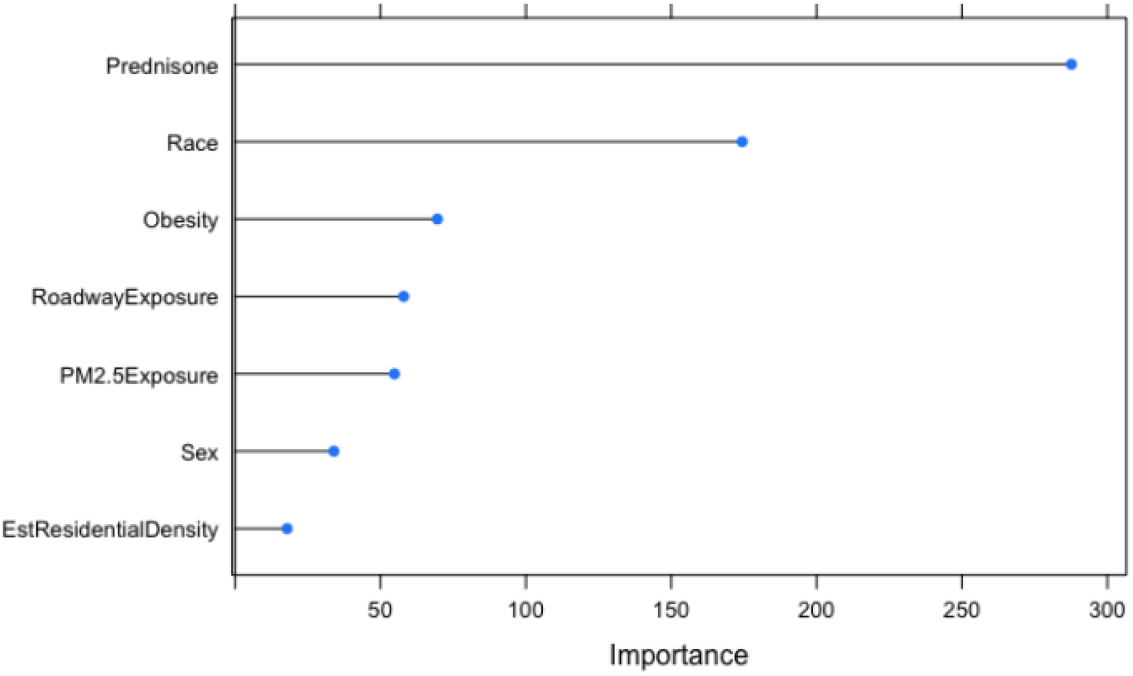
Relative feature importance for all feature with respect to TotalEDInpatientVisits. See Table 1 for feature variable definitions.

### 2.2 Modeling causal networks

Most of the naturally occurring trends that we come across are simply passive observations of events occurring in the world that are either coincidental or unexplained associations. For example, statements like “Drinking beer everyday increase chance of prostate cancer” are common in the news and scientific reporting and in our day-to-day personal beliefs. These associations can be easily mistaken as causation, making us susceptible to logical fallacies without knowing the real underlying cause. Causal inference is the science of learning cause from effect [1]. It is an important field of research because it helps us eradicate spurious correlation [6, 6–8]. The primary aim of inferring causal relations from data is to discover interactions between different entities in the form of *V*_*i*_ → *V*_*j*_, where *V*_*i*_ and *V*_*j*_ are observable features in domain and the arrow indicates that the state of *V*_*i*_ influences the state of *V*_*j*_. Causal inference can be either discovered through observational measurements (seeing) or from measurements after performing some external manipulation/intervention (doing). A causal network [1, 9] can be represented with a directed acyclic graph (DAG) *G* = (*V, E*), where *V* = *V*_*i*_,, *V*_*n*_ denotes the set of features and *E* ∈ (*V* × *V*) denotes the set of edges that are causal in nature. For a causal edge (*V*_*i*_, *V*_*j*_), we say that *V*_*i*_ is a cause (parent) of *V*_*j*_, and *V*_*j*_ is the resulting effect (child) of *V*_*i*_. Let *pa*(*V*_*i*_) denote the set of parents of *V*_*i*_. The conditional probability distribution *P*_*i*_ defines the probability of Vi given the state of its parents *pa*(*V*_*i*_). A causal network represents a joint distribution *P* over variables *V* as long as it satisfies two main assumptions:

a. Causal Markov assumption: Any given variable *V*_*i*_ is independent of its non-descendants, conditioned on all of its direct causes (parents). This implies that the joint distribution *P* (*V*) can be factored as: 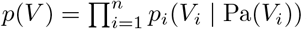.
b. Faithfulness assumption: The joint distribution *p*(*V*_1_, …, *V*_*n*_) is faithful to *G* if every conditional independence relation in the probability distribution *P* is entailed by the Markov assumption applied to *G* [10].

To reconstruct a causal graph from data, we generally start by finding an approximation of the graph, given *V*, and then optimize based on conditions on data. The two main approaches used for causal network inference are:

1. Score-based: This is based on a Bayesian scoring function *S*(*G* | *D*), which estimates the goodness-of-fit of graph *G* to the data *D* [11], as objective functions to maximize, while favoring simpler structures. The score function is usually combined with a search heuristic that explores the space of all possible graphs. Score-based methods are robust and can be extended to include interventional studies (if available), but they are not scalable as network or data size increases.
2. Constraint-based: This method is based on estimating some of the conditional (in)dependencies in the distribution *P* from the data *D* by performing hypothesis tests of conditional independence. Constraint-based methods usually start with a fully connected, undirected graph and progressively remove edges whenever a new conditional independence relation is discovered, while satisfying the corresponding d-separation statements. In this work, we will use a constraint-based approach called the PC algorithm, given that the dataset is observational. To infer the causal graph from data, we learn the equivalence class of a directed acyclic graph (DAG) from data with the traditional constraint-based PC algorithm proposed by [9]. Given a dataset *D* having *n* features *V*_*i*_,, *V*_*n*_, we conduct the following steps. We start with a complete undirected graph given *n* features. We then eliminate edges between variables that are unconditionally independent. For each pair of variables (*V*_*i*_, *V*_*j*_) with an edge between them, and for each variable *V*_*k*_ with an edge connected to either of them, we eliminate the edge between *V*_*i*_ and *V*_*j*_ if *V*_*i*_ ╨ *V*_*j*_ | *V*_*k*_. For each pair of variables *V*_*i*_, *V*_*j*_ having an edge between them, and for each pair of variables *V*_*k*_, *V*_*l*_ with edges both connected to *V*_*i*_ or both connected to *V*_*j*_, we eliminate the edge between *V*_*i*_ and *V*_*j*_ if *V*_*i*_ ╨ *V*_*j*_ | *V*_*k*_, *V*_*l*_. We continue to check independencies conditional on subsets of variables of increasing size *n* until there are no more adjacent pairs (*V*_*i*_, *V*_*j*_) such that there is a subset of variables of size *n* in which all of the variables in the subset are adjacent to *V*_*i*_ or adjacent to *V*_*j*_. For each triple of variables (*V*_*i*_, *V*_*j*_, *V*_*k*_) such that *V*_*i*_ and *V*_*j*_ are adjacent, *V*_*j*_ and *V*_*k*_ are adjacent, and *V*_*i*_ and *V*_*k*_ are not adjacent, we orient the edges *V*_*i*_––*V*_*j*_––*V*_*k*_ as *V*_*i*_ → *V*_*j*_ ← *V*_*k*_, if *V*_*j*_ is not in the set conditioning on which *V*_*i*_ and *V*_*k*_ became independent and the edge between them was accordingly eliminated. We call such a triple of variables a v-structure. For each triple of variables such that *V*_*i*_ → *V*_*j*_––*V*_*k*_, and *V*_*j*_ and *V*_*k*_ are not adjacent, we orient the edge *V*_*j*_––*V*_*k*_ as *V*_*j*_ → *V*_*k*_. This is called orientation propagation.

## 3 Results

### 3.1 Inferring causal graphs

We first applied the PC algorithm to the ICEES multivariate feature table. In Fig. 3a, we show the inferred casual graphs, first using the entire table with all eight features and second in Fig. 3b using only the top four important features with respect to TotalEDInpatientVisits, as determined in section 2.1. Expected relationships between features based on subject matter expertise and published literature are represented in black (solid and dashed) lines. There are eight such expected edges, which we use to measure the structure-learning accuracy of the causal algorithm. Solid black lines represent expected edges (true positives) that are reported via the PC algorithm, while dashed lines are edges which were expected but missed (false negatives). Newly found relationships inferred by the PC algorithm, that are not expected, are represented in red (false positive). We note that there were a few undirected edges detected, for which the algorithm was not able to determine directionality.

**Fig 3:**
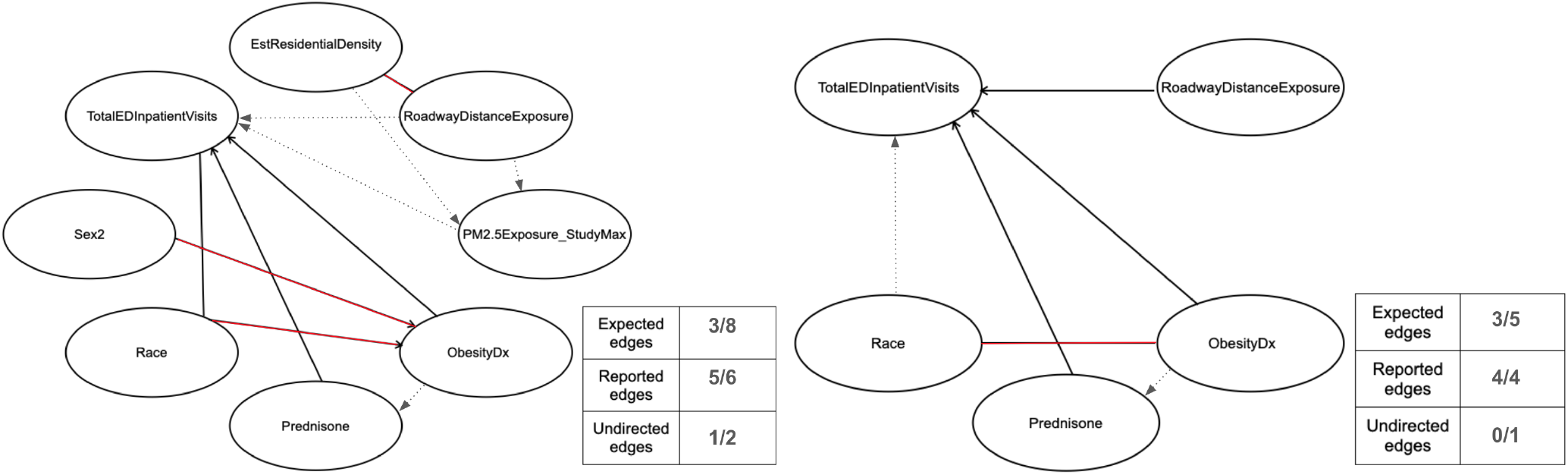
Inferred causal graph. Solid black lines represent true positives, dashed lines represent false negatives and red lines represents false positives

Three of eight expected edges were inferred. For the determined false positive edges, we conducted a further literature survey to find multiple (5+) citations where a relationship between the features were reported. We marked them as reported edges, along with the true positives. Two out of the three additional edges detected were found in the literature; hence, we marked them as reported. The expected directed edge from Race → TotalEDInpatientVisits was also missed.

As discussed in section 2, we queried the openAPI a second time to generate a multivariate table containing only the top four important features (Prednisone, Race, Obesity and RoadwayExposure) with respect to TotalEDInpatientVisits, as identified by random forest (Fig. 3b). We found significant improvement in accuracy. Three out of five expected edges were detected. An undirected edge between Race and ObesityDx was also detected, which are reported in literature as highly associated features.

### 3.2 Effects of Intervention

Having learned a causal network from the data, we now use it to answer relevant questions by making inferences. To evaluate this, we computed the effects of interventions on features by modifying the network to simulate interventions. Firstly, because some of the edges detected in Fig. 3a were undirected, we removed them. We then learned the parameters of our learned causal DAG given the network structure and the data. Next, we constructed a mutilated network to simulate a perfect intervention by setting a target node to a particular value. Finally, we tested the effects of these interventions, while verifying the correctness of the learned causal network, to substantiate some commonly known causal links like the following expected claims:

- **Claim (a)**. Obesity should have a direct effect on TotalEDInpatientVisits. Hence, conducting an intervention on the node “Obesity” should reflect a change (increase or decrease, accordingly) in the probability distribution of TotalEDInpatientVisits.
- **Claim (b)**. Prednisone should have a direct effect on TotalEDInpatientVisits. Hence, conducting an intervention on the node “Prednisone” should reflect a change (increase or decrease, accordingly) in the probability distribution of TotalEDInpatientVisits.
- **Claim (c)**. Sex2 should not have a direct effect on TotalEDInpatientVisits. Hence, conducting an intervention on the node “Sex2” should not reflect a change (increase or decrease, accordingly) in the probability distribution of TotalEDInpatientVisits.

We conducted these three interventions on our learned causal network. To test Claim (a), we created a mutilated network by fixing the state of ObesityDx to 1, which means we are forcing ObesityDx to be present. For Claim (b), we fixed the state of Prednisone to be 1, again meaning that we are forcing prednisone to be present. For Claim (c), we fixed state of Sex2 to be Male. Next, we compared the changes in the probability distribution of TotalEDInpatientVisits before and after these three *ad hoc* interventions to confirm the expected causal influences. We plotted the changes in the probability distribution of TotalEDInpatientVisits in Fig. 4. As expected, there were changes in the probability distribution of TotalEDInpatientVisits for interventions a and b, reflected in Fig. 4a and b, respectively. For intervention c, the changes before and after intervention were negligible, meaning that Sex2 had no causal effect on the frequency of TotalEDInpatientVisits.

**Fig 4:**
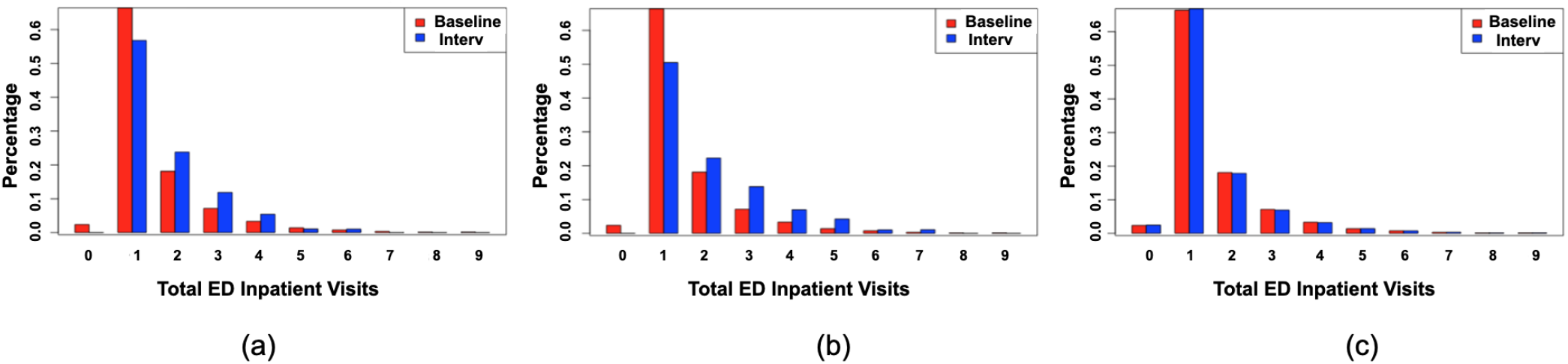
Effect of intervention on (a) Obesity, (b) Prednisone and (c) Sex: change in the probability distribution of TotalEDInpatientVisits before (red) and after (blue) intervention.

## 4 Discussion

We demonstrated the ability to use the ICEES OpenAPI to answer important questions about causal relationships between factors affecting asthma attacks. We focused on a large cohort of patients with asthma or related conditions and a dataset that included data derived from EHRs and a variety of public sources of environmental exposures data. We applied PC analysis, a constraint-based causal learning algorithm, on the dataset and identified prednisone, race, and obesity as significant predictors of annual ED or inpatient visits for respiratory issues, followed by residential distance from a major roadway/highway, airborne particulate exposure, and sex. Of those, prednisone and obesity were found to be causally related to annual ED or inpatient visits in our causal inference model, and sex and race were found to be indirectly related to annual ED or inpatient visits via a causal relationship to obesity. On a smaller dataset, comprising only the four most important features, as determined by random forest analysis, we identified residential distance from a major roadway/highway as an additional variable that is casually related to annual ED or inpatient visits for respiratory issues.

We validated our findings based on expert knowledge and prior published literature. Most of our results are consistent with previously published literature [12]. For instance, prednisone, which is commonly prescribed for patients who are non-responsive to first-line treatments such as inhaled albuterol [13], has been identified as a factor associated with asthma exacerbations and ED or inpatient visits for respiratory issues [14]. Female sex, obesity, and African American race have previously been identified as factors that contribute to asthma attacks [15]. In another work by our group [4] and others [16], obesity and sex have been found to be highly related to asthma attacks. Several other works [17, 18] have additionally found a significant association between African American race and increased risk of asthma attacks. Exposure to major roadways or highways has also been found to be a risk factor for asthma. Several studies [19, 20] have demonstrated an increase in asthma attacks among patients residing in close proximity to a major roadway or highway. Our findings on the relationship between roadway exposures and asthma exacerbations have been inconsistent, with evidence to support [14] and negate [12] a relationship.

One factor that we expected to find in our model as causally related to asthma attacks, but did not, is exposure to airborne particulate matter. Exposure to airborne particulate matter is a well-established trigger for asthma attacks [3, 12, 14, 15, 21]. The failure to detect a causal relationship between exposure to airborne particulate matter and asthma attacks likely reflects the imbalance in the distribution of patients across bins. Indeed, we are actively refining both our exposure models and our binning strategy. For instance, instead of using a Python algorithm to bin the airborne pollutant exposures, we are considering a binning strategy based on subject matter expertise.

## 5 Conclusion

EHR data, while being rich data sources for important clinical information, are mostly observational and generally challenging to access due to regulatory constraints. Performing real-world interventions are not only costly, but even impractical, given the need to integrate large data sources across various domains. Causal inference provides an excellent tool to simulate clinical interventions and answer questions about the effects of medical and healthcare interventions. In this study, we used the regulatory-compliant open ICEES service to generate a multivariate feature table and apply a causal inference model, as well as conduct simulated interventions, to explore the influence of key demographic factors and environmental exposures on asthma attacks. Our results were largely consistent with expectations based on subject matter expert opinion and published literature. As part of our future studies, we are expanding our causal inference modeling to include additional features and additional years of data in order to reflect the underlying causal relationships at a larger scale, while supporting additional use cases, including primarily ciliary dyskinesia and other rare respiratory disorders.

### 5.1 Availability

The ICEES asthma OpenAPI can be accessed at https:icees-asthma.renci.org/apidocs. The associated public GitHub repositories include: https://github.com/ExposuresProvider/icees-api; https://github.com/NCATS-Tangerine/FHIR-PIT; https://github.com/NCTraCSIDSci/camp-fhir.

## Data Availability

All data produced in the present work are contained in the manuscript

https://icees-asthma.renci.org/apidocs

## 6 Acknowledgements

The authors wish to acknowledge Stanley C. Ahalt, Director of the Renaissance Computing Institute, for support and advice on the work described herein; David B. Peden for his expertise on the asthma use case; Emily R. Pfaff and James Champion for their help with the patient data; and Sarav Arunachalam, Stephen A. Appold, Alejandro Valencia Arias, and Lisa Stillwell for their help with the environmental exposures data.

## References

1. Judea Pearl. Causality: models, reasoning, and inference. Econometric Theory, 19(675-685):46, 2003.

2. Dominick A Rizzi. Causal reasoning and the diagnostic process. Theoretical medicine, 15(3):315–333, 1994.

3. Karamarie Fecho, Emily Pfaff, Hao Xu, James Champion, Steve Cox, Lisa Stillwell, David B Peden, Chris Bizon, Ashok Krishnamurthy, Alexander Tropsha, et al. A novel approach for exposing and sharing clinical data: the translator integrated clinical and environmental exposures service. Journal of the American Medical Informatics Association, 26(10):1064–1073, 2019.

4. Karamarie Fecho, Stanley C Ahalt, Saravanan Arunachalam, James Champion, Christopher G Chute, Sarah Davis, Kenneth Gersing, Gustavo Glusman, Jennifer Hadlock, Jewel Lee, et al. Sex, obesity, diabetes, and exposure to particulate matter among patients with severe asthma: Scientific insights from a comparative analysis of open clinical data sources during a five-day hackathon. Journal of biomedical informatics, 100:103325, 2019.

5. Max Kuhn. Building predictive models in r using the caret package. Journal of statistical software, 28(1):1–26, 2008.

6. Meghamala Sinha. Causal structure learning from experiments and observations. 2019.

7. Meghamala Sinha, Prasad Tadepalli, and Stephen A Ramsey. Pooling vs voting: An empirical study of learning causal structures. 2019.

8. Meghamala Sinha, Prasad Tadepalli, and Stephen A Ramsey. Voting-based integration algorithm improves causal network learning from interventional and observational data: an application to cell signaling network inference. Plos one, 16(2):e0245776, 2021.

9. Peter Spirtes, Clark Glymour, and Richard Scheines. Causation, prediction, and search. Adaptive computation and machine learning. MIT Press, Cambridge, MA, 2000.

10. Marek J Druzdzel. The role of assumptions in causal discovery. 2009.

11. Judea Pearl. Graphical models for probabilistic and causal reasoning. In Quantified representation of uncertainty and imprecision, pages 367–389. Springer, 1998.

12. Karamarie Fecho, Perry Haaland, Ashok Krishnamurthy, Bo Lan, Stephen A Ramsey, Patrick L Schmitt, Priya Sharma, Meghamala Sinha, and Hao Xu. An approach for open multivariate analysis of integrated clinical and environmental exposures data. Informatics in Medicine Unlocked, 26:100733, 2021.

13. Abdullah A Alangari. Corticosteroids in the treatment of acute asthma. Annals of thoracic medicine, 9(4):187, 2014.

14. Karamarie Fecho, Stanley C Ahalt, Stephen Appold, Saravanan Arunachalam, Emily Pfaff, Lisa Stillwell, Alejandro Valencia, Hao Xu, David B Peden, et al. Development and application of an open tool for sharing and analyzing integrated clinical and environmental exposures data: Asthma use case. JMIR Formative Research, 6(4):e32357, 2022.

15. Bo Lan, Perry Haaland, Ashok Krishnamurthy, David B Peden, Patrick L Schmitt, Priya Sharma, Meghamala Sinha, Hao Xu, and Karamarie Fecho. Open application of statistical and machine learning models to explore the impact of environmental exposures on health and disease: An asthma use case. International Journal of Environmental Research and Public Health, 18(21):11398, 2021.

16. Rebecca E Greenblatt, Edward J Zhao, Sarah E Henrickson, Andrea J Apter, Rebecca A Hubbard, and Blanca E Himes. Factors associated with exacerbations among adults with asthma according to electronic health record data. Asthma research and practice, 5(1):1–11, 2019.

17. Hao Xu, Steven Cox, Lisa Stillwell, Emily Pfaff, James Champion, Stanley C Ahalt, and Karamarie Fecho. Fhir pit: an open software application for spatiotemporal integration of clinical data and environmental exposures data. BMC medical informatics and decision making, 20(1):1–8, 2020.

18. Corinne A Keet, Meredith C McCormack, Craig E Pollack, Roger D Peng, Emily McGowan, and Elizabeth C Matsui. Neighborhood poverty, urban residence, race/ethnicity, and asthma: rethinking the inner-city asthma epidemic. Journal of Allergy and Clinical Immunology, 135(3):655–662, 2015.

19. Laura Perez, Fred Lurmann, John Wilson, Manuel Pastor, Sylvia J Brandt, Nino Künzli, and Rob McConnell. Near-roadway pollution and childhood asthma: implications for developing “win–win” compact urban development and clean vehicle strategies. Environmental health perspectives, 120(11):1619–1626, 2012.

20. Shepherd H Schurman, Mercedes A Bravo, Cynthia L Innes, W Braxton Jackson, John A McGrath, Marie Lynn Miranda, and Stavros Garantziotis. Toll-like receptor 4 pathway polymorphisms interact with pollution to influence asthma diagnosis and severity. Scientific reports, 8(1):1–11, 2018.

21. Maria C Mirabelli, Ambarish Vaidyanathan, W Dana Flanders, Xiaoting Qin, and Paul Garbe. Outdoor pm2. 5, ambient air temperature, and asthma symptoms in the past 14 days among adults with active asthma. Environmental health perspectives, 124(12):1882–1890, 2016.

